# Metabolic alterations in Crohn’s disease: A Systematic Review

**DOI:** 10.1101/2024.10.03.24314812

**Authors:** Atul Dev, Sartajdeep Kahlon, Jonathan Javier Ruiz, Avinash Chandra Kushwaha, Megan G. Van Noord, Sean H. Adams, Kim Elaine Barrett, Adam Paul Arkin, Maneesh Dave

## Abstract

**Background:** Crohn’s disease (CD) is a chronic inflammatory disorder of the gastrointestinal tract with an unknown etiology. Several studies have identified dysregulated metabolites in patients with CD. However, there is significant variability in the metabolites found to be dysregulated across these studies, making it unclear whether a comprehensive, disease-specific metabolic signature for CD exists.

**Objective:** To analyze Crohn’s disease-specific metabolomic studies and available datasets to identify a comprehensive signature of dysregulated metabolites and metabolic pathways implicated in human CD.

**Design:** A comprehensive systematic review was conducted using Medline and Embase databases to identify studies (from inception to May 2024) that employed analytical chemistry techniques to quantify metabolites in various biological samples from Crohn’s disease patients and non-IBD controls. Metabolites that were significantly altered in Crohn’s patients and reported in at least two studies were included for further analysis.

**Results:** The systematic search identified 3,632 studies, with 88 selected for data extraction. Across these studies, 79 metabolites were consistently reported as significantly altered in Crohn’s disease (CD) patients in two or more studies. These metabolites form a distinct metabolic signature that differentiates CD patients from non-IBD controls, highlighting their relevance in the pathophysiology of the disease.

**Conclusion:** This systematic review presents a comprehensive and well-defined signature of dysregulated metabolites across various biological samples and provides detailed insight into the perturbed metabolic pathways involved in CD.

## 1. Introduction

Inflammatory bowel disease (IBD), consisting of Crohn’s disease (CD) and ulcerative colitis (UC), is a chronic relapsing-remitting inflammatory disorder of the gastrointestinal (GI) tract.[1] CD is characterized by transmural inflammation along any part of the digestive tract, which may lead to complications such as abscesses, fibrotic strictures, or fistulas.[2] The pathogenesis of CD involves epithelial barrier dysfunction, immune dysregulation, and gut dysbiosis.[3] These disruptions alter metabolic pathways, which may contribute to the initiation and/or perpetuation of chronic inflammation.[4]

Recent advancement of “omics” technologies has significantly enhanced our understanding of Crohn’s disease (CD) pathophysiology. Among these, metabolomics has emerged as a promising field focused on the systematic identification and quantification of small molecules (metabolites) in biological samples.[5] Each metabolite, with its unique chemistry and function, provides valuable insights into the metabolic processes occurring in the body, offering a window into the molecular and cellular changes that distinguish healthy individuals from those with disease. Current metabolomics platforms utilize both targeted and untargeted approaches, employing a range of high-throughput analytical techniques to detect and characterize metabolites, providing detailed information about their structure and concentration.

As CD is a complex disease, identifying dysregulated metabolites can enhance our understanding of the metabolic perturbations associated with the disease and potentially inform the development of more effective, targeted therapies. Although numerous studies have quantified and compared dysregulated metabolites in IBD patients versus non-IBD controls, significant differences exist due to differences in patient populations, disease location, biological sample types, and detection techniques. This systematic review aims to synthesize and evaluate all reported studies and datasets on dysregulated metabolites in a diverse, global CD population, with the goal of identifying a unique, comprehensive metabolite signature. By compiling these altered metabolites, we aim to provide a detailed overview of the metabolic changes associated with CD, contributing to a more precise molecular understanding of its pathophysiology.

## 2. Methods

We followed the standard PRISMA statement for performing and reporting systematic review.

### 2.1 Search strategy

A comprehensive literature search was conducted using the Medline and Embase databases (from inception till May 2024). The search was independently performed by authors (AD, SK, JJR and ACK) with the help of researcher services librarian (MGVN). Four authors (AD, SK, JJR and ACK) performed initial screening of the studies and resolved any discrepancies through consensus with the senior author (MD). We used free text words and MeSH terms with and without Boolean operators (“AND” and “OR”) to increase the sensitivity of the search **(supplementary table S1).** The in-detailed search strategy, study inclusion and exclusion criteria, data analysis strategy are available in **supplementary information.**

## 3.0 Results and discussion

### 3.1 Search results

A total of 3632 studies were identified prior to excluding the duplicates [n = 91]. For initial screening, title and abstracts of 3541 studies were assessed and irrelevant studies were excluded [n = 3422] if they were not original studies or did not directly reference CD, analytical techniques, metabolomics, or metabolites. A total of 64 studies were resolved by consensus with several studies having been initially excluded by one reviewer as they only referenced UC or IBD but were eventually included as they conducted subgroup analysis of CD patients. In total 119 studies were sought for full text review, among those, few studies were removed [n = 31] for reasons including ‘wrong indication’, where studies did not include metabolites, ‘wrong patient population’, where less than 10 CD patients were included or did not utilize non-CD controls, ‘wrong comparator’ where comparison was not between CD patients and non-IBD controls and where no analytical chemistry techniques was used to quantify the metabolites. Consequently, 88 full text articles comprising in-total of 5658 CD patients and 5375 non-IBD controls were included as a part of the review. A PRISMA flow diagram associated with this search is shown in **Figure 1**.

**Figure 1.**
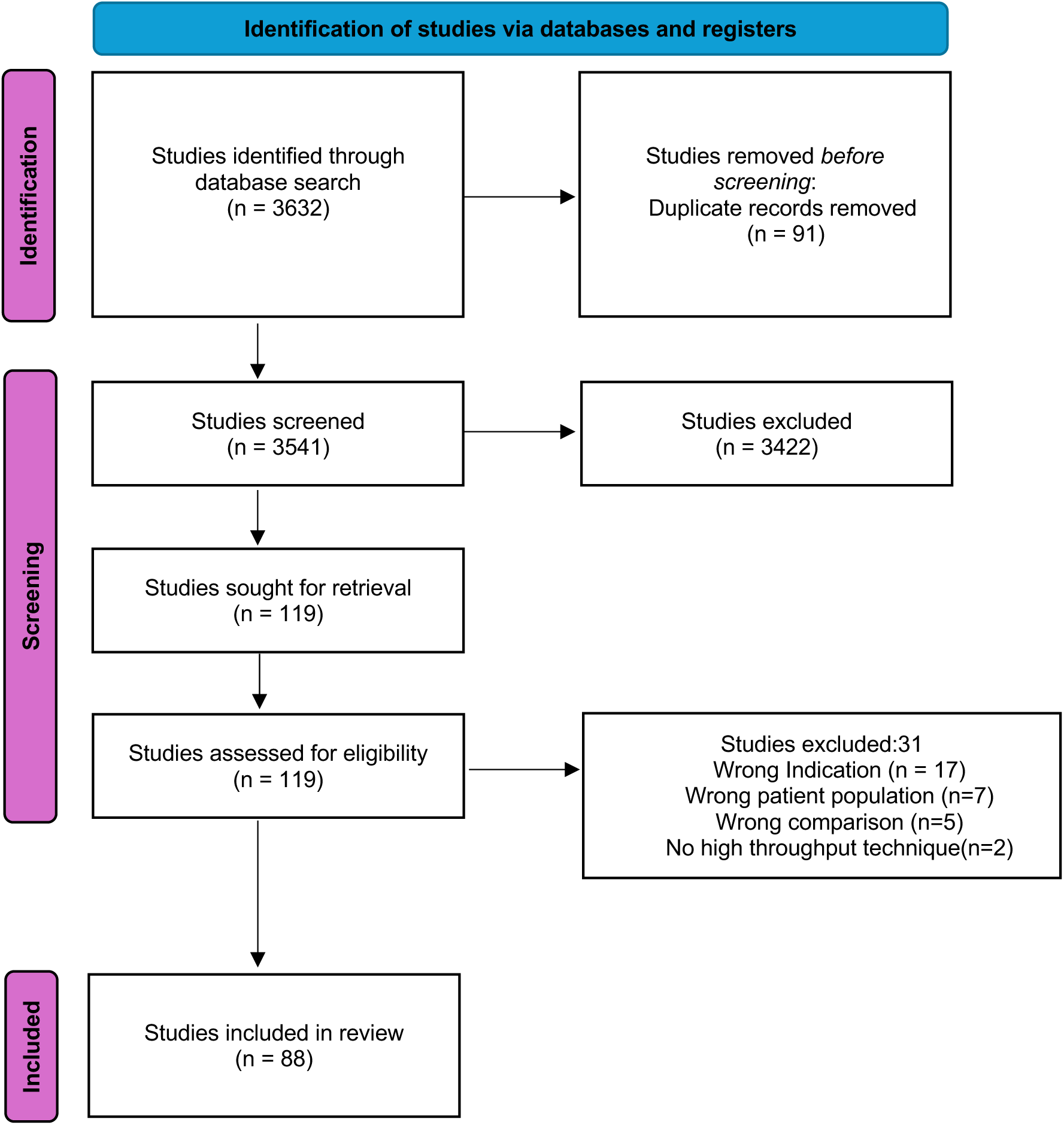
Preferred Reporting Items for Systematic Reviews and Meta-Analyses (PRISMA)

### 3.2 Dysregulated metabolites in Crohn’s disease

Metabolites play an important role in maintaining homeostasis in the human body. Prolonged disruptions in metabolic pathways can lead to symptomatic phenotypes, with a disease-specific signatures in biological tissues, that can be quantified using metabolomic analytical techniques. **(Figure S1).** In the following paragraphs we described metabolic perturbations in CD in the four major classes.

#### 3.2.1 Lipids and Fatty acids

Fats, a combination of triglycerides (TGs) and fatty acids (FAs) are crucial components of cellular membranes and play vital roles in energy storage, signaling, and structural integrity of the cells. Alterations in these metabolites can disrupt biological processes, potentially leading to pathological conditions such as CD. In our systematic review, we observed SCFAs abnormalities in multiple biological samples of CD patients. Levels of butyrate/butyric acids were consistently low in the stool samples; however, no consistent pattern was observed for acetate, propionate and valerate. Butyrate is synthesized via breakdown of dietary fibers by microbiota present in the colon and serves as the main energy source for colonocytes, meeting approximately 70% of their daily energy demand.[6, 7] CD is characterized by gut dysbiosis, with studies showing that butyrate-producing bacteria, such as *Faecalibacterium prausnitzii* and *Roseburia hominis,* are significantly lower in CD patients,[8] and are likely responsible for the consistently low butyrate levels observed in their stool samples. Butyrate maintains low oxygen levels in human colon, reduced butyric acid in stool increases oxygen concentration in the lumen, and promotes aerobic bacteria overgrowth further contributing to gut dysbiosis and contribute to activation of immune cells via multiple inflammatory mechanism **(Figure S2)**.[9] In an attempt to restore the lost microbial diversity of butyrate producing bacteria in CD patients and to reverse mucosal inflammation, Vaughn et al.[10] performed a prospective open-label study of fecal microbiota transplant (FMT) from healthy controls. In this open-label study FMT was overall safe and expanded the existing microbial bacterial diversity in active CD patients, although, there was variable response in clinical symptoms of CD, indicating that there are multiple contributing factors which governs the disease etiology. In addition to dysregulated levels of butyrate in stool, 2-hydroxybutyrate (2HB), which is non-conventional form of SCFAs and considered as ketone body, showed consistent elevated levels in serum samples of CD patients. Humans produce high levels of ketone bodies like 2HB during increased oxidative and metabolic stress or during increased energy demand in inflammatory disease like CD. Another potential reason for increased levels of 2HB in the serum is the contribution from the microbial metabolism in human gut. In a recent study, Qin et al [11] showed that human gut microbiota produces 2HB from methionine, threonine, aspartic acid and 2aminobutyric acid metabolism. In our systematic review, we observed elevated levels of methionine and threonine in the biopsy samples of CD patients [12, 13] and abundance of *Bacteroidetes* is well known in CD, therefore, providing a possible explanation of increased serum levels of 2HB in CD patients.

In addition to SCFAs, levels of mono-unsaturated fatty acids (MUFAs) and poly-unsaturated fatty acids (PUFAs) were consistently dysregulated in serum samples of CD patients; MUFAs (Oleic acid and Palmitoleic acid) were consistently elevated, while PUFAs, omega-3 fatty acids (Eicosapentaenoic acid; EPA, Docosopentanoic acid; DPA, Docosahexaenoic Acid; DHA) and omega-6 fatty acid (Arachidonic Acid; AA) were consistently low. Elevated levels of oleic acid, palmitoleic acid, linoleic acid (LA) in serum samples of CD patients could be a sign of preferential consumption of fat rich diet or presence of genetic polymorphism in genes responsible for the FAs metabolism. Several studies have shown the presence of genetic polymorphism in PUFA metabolism in CD.[14, 15] The *FADS2* gene which catalyze the conversion of LA and ALA (Alpha linolenic acid) into longer chain omega-6 and omega-3 PUFAs (EPA, DHA and AA) is often dysregulated in CD.[16] In a recent study, mesenteric tissues obtained during ileocolonic resection from CD patients showed decreased expression levels of *FADS2* compared to the healthy controls.[17] The low expression of *FADS2* gene is one potential reason for the consistent low levels of EPA, DHA, and AA in CD patients **(Table 2)** and Dietary restrictions and malabsorption due to intestinal damage could further impair the intake and absorption of essential FAs required for the synthesis of EPA, DPA, DHA and AA, increased demand in inflamed tissues in CD result in its further depletion from the serum. Further, epidemiologic studies have linked increased intake of omega-3 fatty acids with a reduced risk of CD,[18] while, increased intake of omega-6 were linked to high incidence of CD.[19] [20, 21]. Omega-3 and Omega-6 fatty acids show competitive binding for the same enzyme complex, and depending on preferential binding, result in a cascade of pro-inflammatory or anti-inflammatory factors.[22] Therefore, a ratio of omega-3 and omega-6 PUFA is considered as a more significant measure to predict the course of disease rather than individual levels of omega-3 and omega-6 PUFA. A higher omega-3: omega-6 ratio is associated with decreased susceptibility in pediatric CD.[15] Further, there was a consistent dysregulation at the levels of sphingolipids and ceramides in CD patients. Sphingolipids are one of the main building blocks of eukaryotic cell membranes, consisting of a sphingoid backbone that is N-acylated with various FAs to form several ceramide species.[23] As a class, sphingolipid and ceramides were elevated in multiple biological samples in CD. However, different studies have measured different sphingolipids and ceramides (varying the carbon chain length; C16, C18 etc.) with little overlap in structure. Sphingolipids influence the function and trafficking of immune cells by interacting with the cell surface receptors and intracellular signaling molecules. [23] Ceramides, a central component of sphingolipid metabolism, can activate various stress signaling pathways, to produce pro-inflammatory cytokines TNF-α and IL-1β.[24] Furthermore, the dysregulation of sphingolipid and ceramide metabolism in CD could exacerbates intestinal barrier dysfunction, increasing permeability and facilitating the translocation of bacteria and other antigens, which in turn perpetuates the inflammatory cycle **(Figure S3).** A detailed list of dysregulated lipids and FAs in the biological samples from CD patients and non-IBD controls is provided in **Table 1**.

**Table 1.**
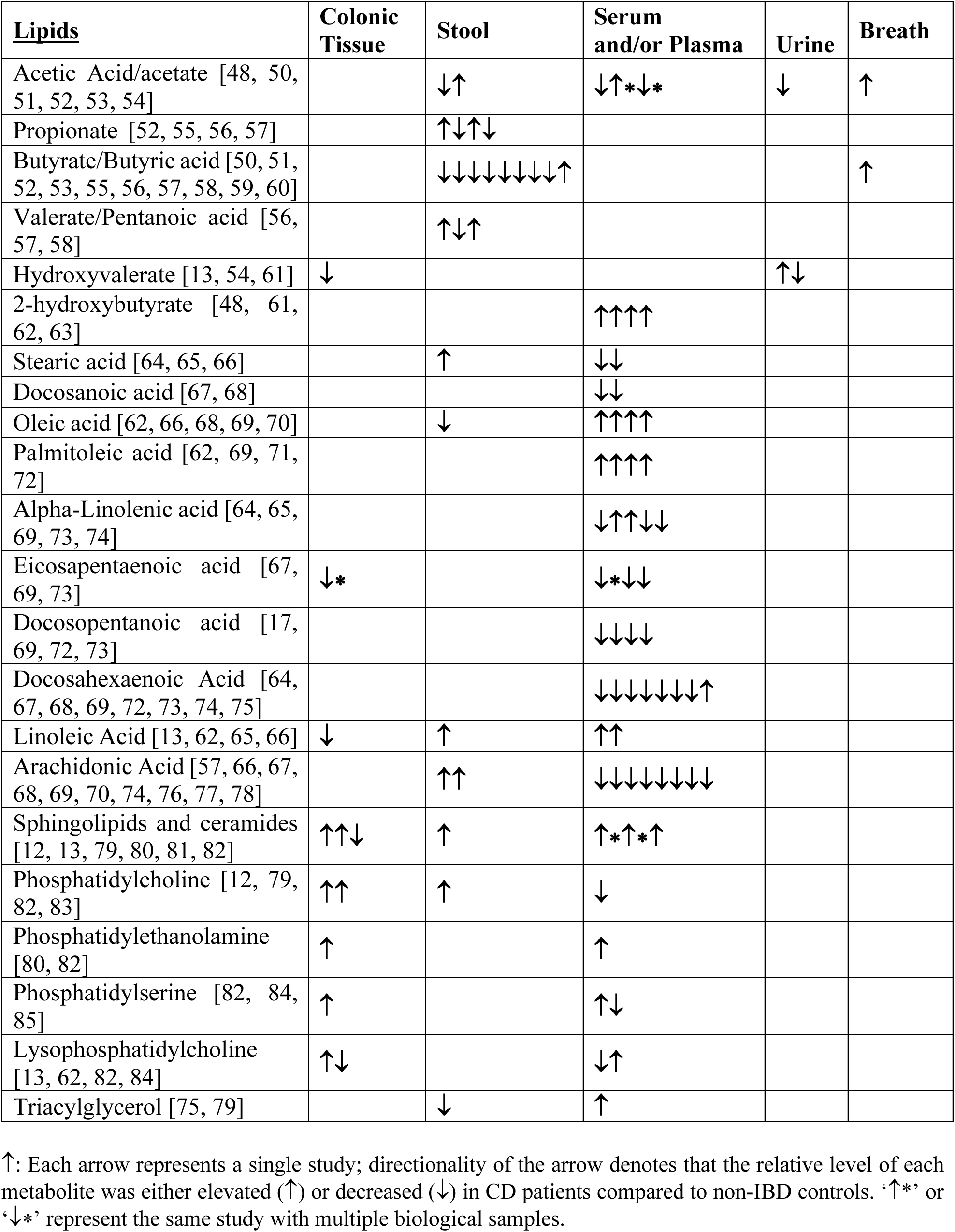
Relative concentrations of fatty acids and annotated lipid metabolites in CD compared to non-CD controls.

**Table 2.**
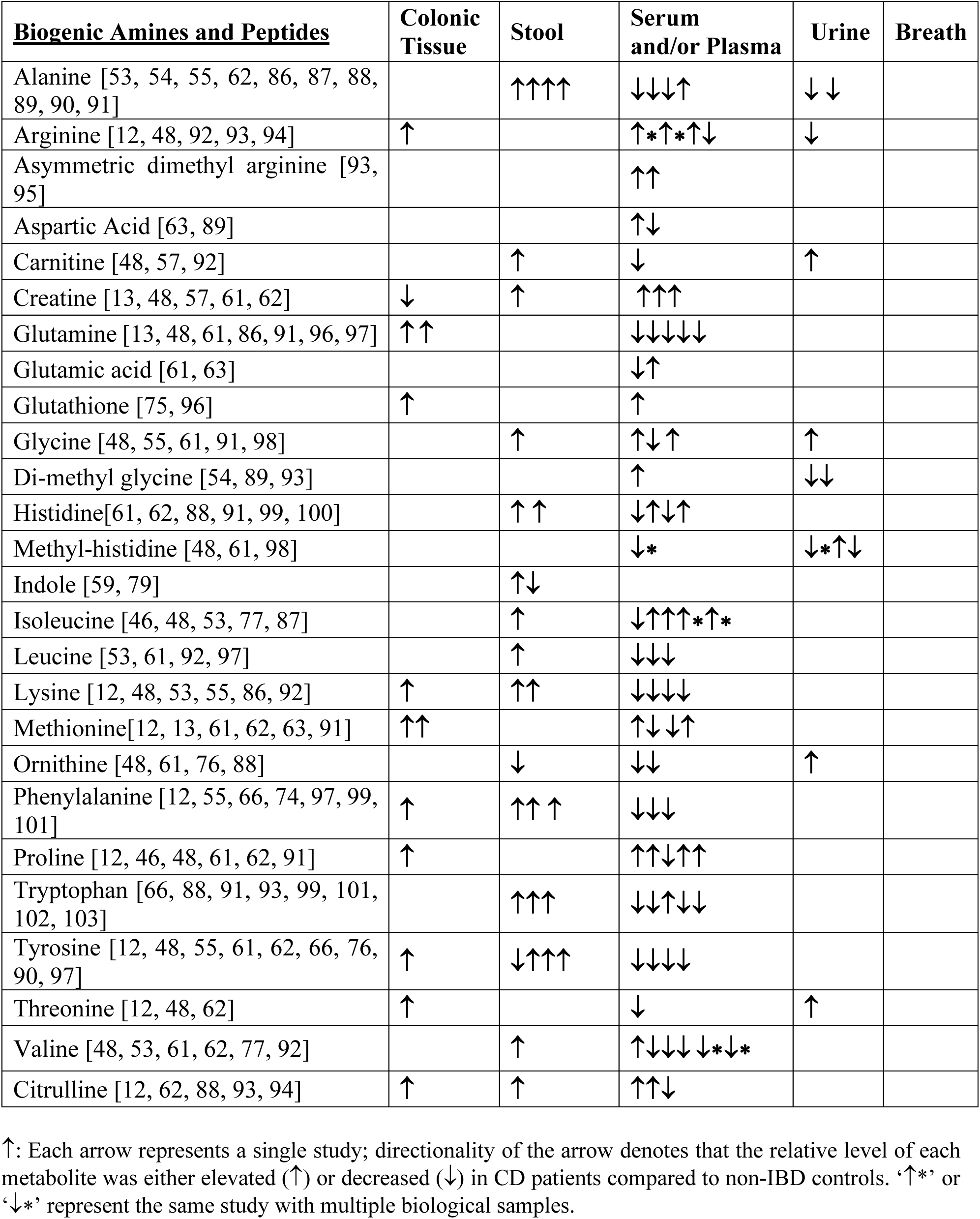
Relative concentrations of biogenic amines and peptides in CD compared to non-CD controls.

#### 3.2.1 Biogenic Amines and Peptides

A biogenic amine is a biologically synthesized molecule with one or more amine groups. Amino acids (AAs) and the proteins are some of the most common examples of biogenic amines in the human body. In our systematic review, we observed dysregulation in the serum levels of essential amino acids (EAAs) and non-essential amino acids (NEAAs) in the CD patients. Five of nine EAAs, including leucine (Leu), lysine (Lys), phenylalanine (Phe), valine (Val) and tryptophan (Trp) were consistently low. Similarly, the levels of NEAAs including, tyrosine (Tyr), alanine (Ala), and glutamine (Gln) followed a similar pattern in the serum and plasma samples of CD patients. In contrast, elevated levels of EAAs Isoleucine (Ile) and the NEAAs Proline (Pro), arginine (Arg) were observed in the serum. The low levels of EAAs could result from intestinal damage caused by chronic inflammation in CD, which impairs nutrient absorption and reduces their availability in serum. Abnormality in NEAAs. which are primarily synthesized by the human body, indicate compromised AAs metabolism in CD patients. Additionally, active inflammation leads to elevated levels of inflammatory cytokines like TNF-α, IL-6, and IL-1β which promote protein catabolism[25] to meet the increased demand for AAs by the immune cells, [26] causing rapid depletion of certain preferential AAs from the serum.

Glutamine (Gln) and Ala are two of the most preferential fuel sources for immune cells, supporting rapid proliferation and cytokine production. Additionally, gut cells utilize Gln to promote enterocyte proliferation, regulate tight junctions, and protect cells from apoptosis and stress under normal and pathological conditions[27]. Consistent low serum levels of Gln and Ala in CD patients suggest chronic inflammation and a high immune cell demand , potentially leading to their depletion in plasma and serum. [28] Interestingly, despite low serum levels, Ala was noted to be highly elevated in the stool samples of CD patients. Ala is a microbial metabolite produced by *Proteobacteria* in the human gut. A recent microbiota characterization study of IBD patients in Denmark by Andersen et al [29] showed a significant increase in the *Proteobacterium* phylum in CD patients, potentially explaining the elevated levels of Ala in stool samples of CD patients. In a separate category of AAs, Leucine (Leu), and Valine (Val) known as branched chain amino acids (BCAAs), play an important role in the regular function of human immune cells and the maintenance of muscle mass.[30] These BCAAs were depleted in the serum samples of CD patients, while levels of another BCAA isoleucine (Ile) were consistently elevated. The gut dysbiosis associated with CD can result in imbalance in the Ile metabolizing bacteria, which may lead to its preferential synthesis or poor catabolism, contributing to elevated levels in serum. While the changes in BCAAs levels have been well reported in several diseases like diabetes, however their role in CD, particularly regarding their serum and plasma levels and its clinical implications remain less clear. It is plausible that the dysregulation of BCAAs may reflect chronic inflammation, elevated mitochondrial oxidation, and altered gut microbiota.

Further, aromatic AAs such as Trp, Phe and Tyr levels were consistently low in the serum and plasma samples of CD patients, while highly elevated in stool samples. Enterocytes absorb and transport Trp via B^0^AT1 (encoded by *SLC6A19*) and TAT1 transporters (encoded by *SLC16A10*). The *SLC6A19* transporter expressed primarily in small intestine, with lower levels in the colon. A previous study showed *SLC6A19* and *SLC16A10* gene expression were significantly downregulated in colon and ileal biopsies of active UC and CD patients respectively.[31] This downregulation could be linked to the decreased absorption of Trp, resulting in low serum levels. Consequently, reduced absorption of Trp allows excessive excretion in the stool, causing elevated levels in CD patients. In addition to Trp metabolism alterations, we observed dysregulation in the Tyr and Phe metabolic pathways in CD patients. The low serum levels of Phe and Tyr may be attributed to the disruptions in several key metabolic pathways, including the conversion of Phe to Tyr, catecholamine synthesis, and protein metabolism. Malabsorption may further contribute to their poor absorption and increased stool levels, while increased metabolic demands in CD could reduce Phe levels, subsequently leading to decreased Tyr levels in the serum.[32, 33] In addition to aromatic AAs, levels of creatine, an endogenous AA, are significantly elevated in the serum and plasma samples of CD patients compared to non-IBD controls. Human body synthesizes creatine from methionine, arginine, and glycine and uses it as an energy source for muscles. Higher serum creatine levels indicate increased energy requirement in the muscles due to metabolic stress and inflammation. Other AAs, such as methionine, histidine, aspartic acid, arginine, ornithine, were also dysregulated in CD patients; however, we did not find any consistent trend in their dysregulation. A complete list of dysregulated biogenic amines and AAs in biological samples from CD patients and non-IBD controls is provided in **Table 2**.

#### 3.2.2 Bile acid and bile salts

Bile acids (BAs) are essential components of bile and crucial for the emulsification and absorption of dietary fats and fat-soluble vitamins. BAs are synthesized in the liver from cholesterol and are termed as primary BAs (cholic acid; CA and chenodeoxycholic acid; CDCA). The human body maintains a pool of BAs in enterohepatic circulation, which is primarily (>90%) composed of conjugated CA, CDCA and deoxycholic acid (DCA); with lithocholic acid (LCA), and ursodeoxycholic acid (UDCA) present in traces amount. Approximately 95% of intestinal BAs are reabsorbed in the terminal ileum via the apical sodium-dependent bile acid transporter (ASBT) and by passive diffusion into the hepatic portal vein, with residual ∼5% BAs pool excreted through the feces.[34] The gut microbiota plays a central role in the maintaining the of BAs pool through enzymatic deconjugation and dehydroxylation reactions, producing unconjugated primary and secondary BAs. The gut microbiome deconjugates primary BAs via bile salt hydrolase (BSH); while 7α/β-dehydroxylases and hydroxysteroid dehydrogenases (HSDHs) are responsible for dehydroxylation and epimerization into secondary BAs.[35] B*acteroides spp*. play a major role in deconjugation of primary BAs, while *Firmicutes* are responsible for the 7α/β-dehydroxylation.[36] Gut dysbiosis, common in CD, is characterized by an abundance of *Bacteroides* [4, 37] and loss in diversity of *Firmicutes.*[38] Dysregulation of these microbiota strongly affects the BAs homeostasis in human gut, resulting in altered BAs profile (low serum levels of Taurocholic acid; TCA, Glycholic acid; GCA, Taurochenodeoxycholic acid; TCDCA, Glycochenodeoxycholic acid; GCDCA, Taurodeoxycholic acid;TDCA, Glycodeoxycholic acid; GDCA, Glycolithocholic acid; GLCA, Tauroursodeoxycholic acid; TUDCA) and low stool levels of DCA, LCA, and UDCA and BA induced diarrhea in CD patients. A significant percentage of CD patients undergo small bowel resection, which further decreases reabsorption of BAs resulting in deficiency of BAs in CD patients.[39]

In addition to the fat absorption, BAs play important role in immune regulation in human body. BAs modulate the systemic pro-inflammatory/anti-inflammatory balance by binding to farnesoid X receptor (FXR), Takeda G protein-coupled receptor 5 (TGR5), and other non-canonical BAs receptors **(Figure S4)**.[40, 41] FXR is a nuclear transcription factor and highly expressed in the human gut, liver and innate immune cells such as macrophages.[42] FXR activation in myeloid cells suppresses intestinal inflammation by inhibiting NF-κB/NLRP3-induced expression of TNFα, IL-1β, and IL-6.[43] In addition to FXR, TGR5 plays an important role in BAs homeostasis and shaping intestinal immunity. TGR5 is a G-protein coupled receptor present in the cell membranes and is widely expressed in small intestine, colon, adipocytes, and innate immune cells.[44] TGR5 signaling has an anti-inflammatory effect and prevents the production of pro-inflammatory cytokines TNFα and IL-1β,[45] highlighting the importance of TGR-5 signaling in CD. BAs dysregulation can be attributed to gut dysbiosis in CD, and correcting gut dysbiosis represents a potential therapeutic approach for the treatment of CD. In a recent study, Zhou et al. showed that oral gavage treatment with a BAs consortium (BAC; *Clostridium AP sp000509125, Bacteroides ovatus, and Eubacterium limosum*) in mice had protective effects against DSS-induced colitis, suggesting that a rationally designed bacterial consortium could reshape gut microbiome metabolism to treat CD.[46] A complete list of dysregulated BAs and bile salts in biological samples from CD patients and non-IBD controls is provided in **Table 3**.

**Table 3.**
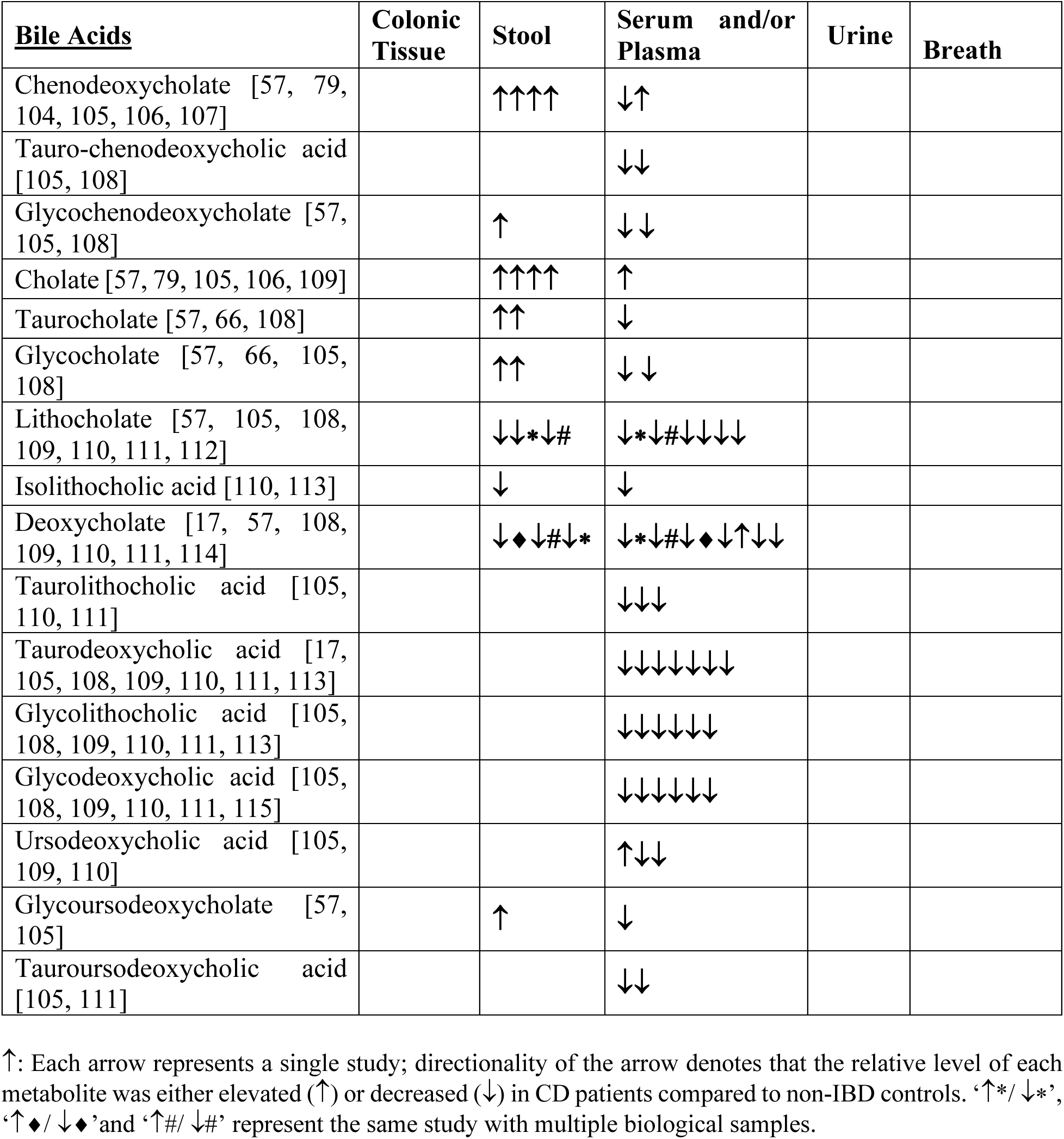
Relative concentrations of bile acids and in CD compared to non-CD controls.

#### 3.2.3 Other primary metabolites

In our systematic review, we observed reduced levels of alpha-ketoglutarate (AKG), succinate, and citrate in the serum and urine samples of CD patients. These metabolites are key intermediates of the TCA cycle, and their dysregulation suggests impaired mitochondrial metabolic pathways. The human body tightly regulates citrate availability and elimination from the body based upon the nutritional intake, cellular metabolism, renal clearance, and bone remodeling. Consistently low levels of citrate in the urine samples of CD patients may be linked to poor absorption from the diet or increased cellular metabolism. CD is characterized by chronic inflammation with increased numbers and activity of immune cells and increased energy demand. To meet these energy requirements, immune cell metabolism shifts towards glycolysis, leading to a compromised TCA cycle. Activated macrophages exhibit a disrupted TCA cycle with reduced isocitrate dehydrogenase (IDH) activity. IDH normally converts citrate to AKG, but its low expression in activated macrophages leads to citrate accumulation in the immune cells and decreased AKG production. Immune cells import citrate both extracellularly and intracellularly to meet their increased energy demands. Accumulated citrate is then metabolized to several intermediates that contribute to the production of pro-inflammatory molecules.[47] Inflammatory environment in CD increase the demand for citrate in the serum to fuel immune cells, resulting in the low levels in urine samples. In addition to citrate, succinate, another metabolite of TCA cycle, is implicated in CD. Succinate shows similarly dysregulated pattern in CD patients compared to non-IBD controls, potentially indicating shared dysregulated pathways. Elevated levels of intracellular and extracellular succinate are pro-inflammatory, contributing to activation and maintenance of the pro-inflammatory state of macrophages during inflammation. Besides, the TCA cycle intermediates, we also observed consistently low levels of hippurate in the urine samples of CD patients. Urinary hippurate is a product of the microbial metabolism of certain dietary compounds, such as benzoic acid, and p-cresol sulphate (a metabolite of *Clostridia* spp. metabolism of tyrosine). In CD, the reduction in Clostridium cluster IV and *Faecalibacterium prausnitzii* has been linked to low levels of hippurate, Schicho et al. [48] reported significantly reduced levels of p-cresol in the urine of CD patients compared to healthy subjects, indicating potential dysbiosis of *Clostridia spp* in CD, which may contribute to the reduced levels of hippurate levels.

In addition to hippurate, several volatile metabolites produced by microbial metabolism and oxidative stress were implicated in CD patients. We detected dysregulated levels of acetate, ethanol, isoprene, pentane, acetone and butanoic acid in breath samples of CD patients compared to non-IBD controls. However, most of these metabolites, except pentane, did not show a consistent pattern in CD. Pentane levels were consistently elevated in breath samples of CD patients, likely as a result of lipid peroxidation due to oxidative stress.[49] CD is characterized by elevated levels of reactive oxygen species and disruptions in lipid metabolism, which may contribute to the increased pentane levels in breath samples. A complete list of other dysregulated primary metabolites in biological samples from CD patients and non-IBD controls is provided in **Table 4**.

**Table 4.**
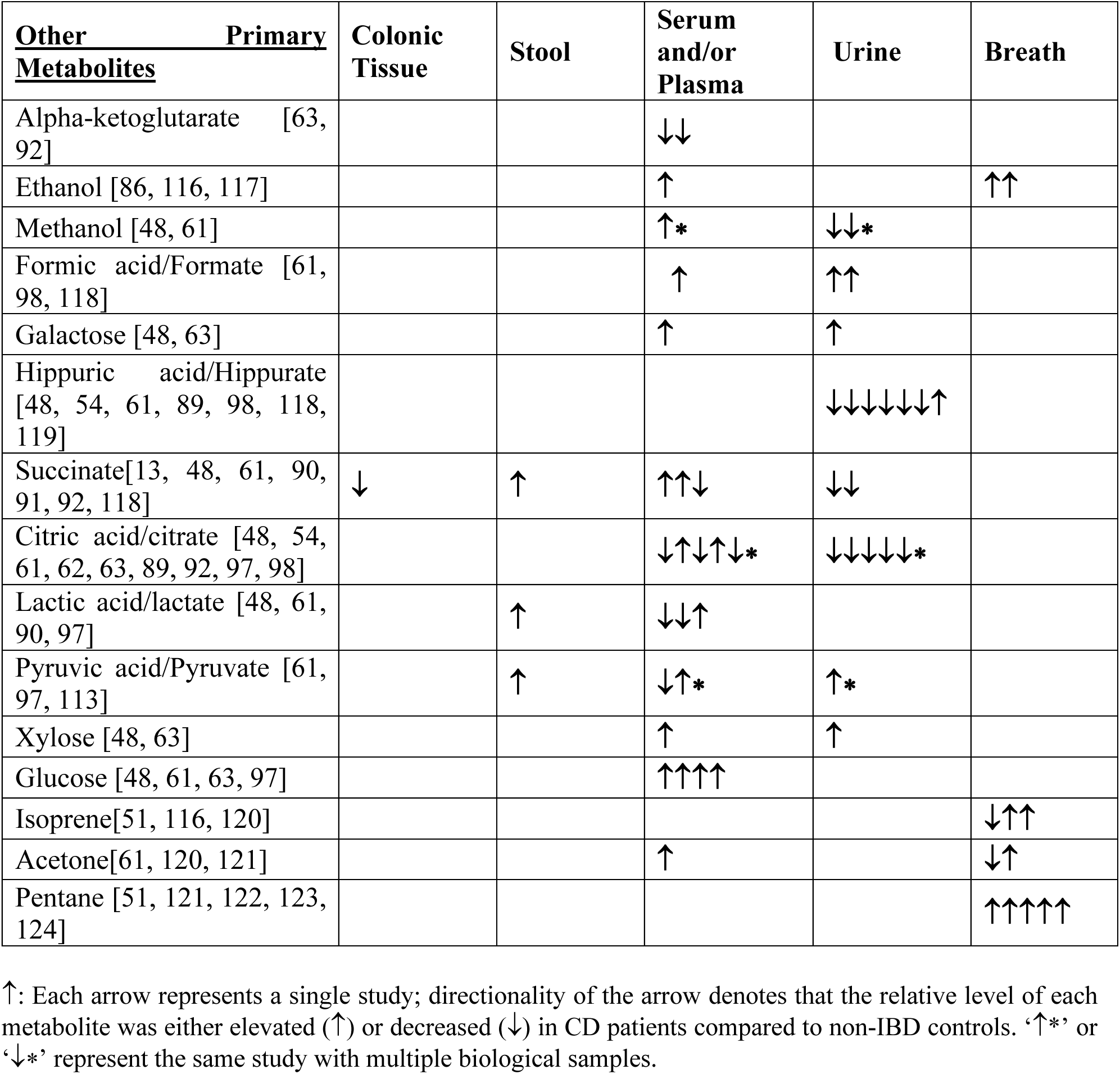
Relative concentrations of other primary metabolites in CD compared to non-CD controls.

## 4. Conclusion

A key takeaway from the systematic review and analyses is that dysregulated metabolites in CD patients compared to non-CD controls, span multiple metabolite classes, including lipids and fatty acids, biogenic amines and peptides, bile acids and bile salts, as well as other primary metabolites. Each category of metabolite provides a distinct signature within the biological samples of CD patients. Notably, certain key metabolites such as butyrate, 2HB, Ile, Ala, primary and secondary BAs, and hippurate, were strongly influenced by the gut dysbiosis linked to the CD. However, it remains unclear whether these dysregulated metabolites are a cause or a consequence of gut dysbiosis in CD. Another strength of our systematic review lies in the comprehensive comparison of individual metabolites between CD patients and non-IBD controls. The metabolites reported were not only consistently identified across multiple studies but were also detected using diverse, high-throughput analytical techniques across various biological sample types, adding robustness to our findings. This study’s limitations include the focus on metabolites identified in at least two independent studies, which may have excluded CD-associated metabolites found in single studies due to differing analytical libraries and resources. Despite this, the metabolites presented are robust and provide a foundation for further exploration of their roles in the CD phenotype. Additionally, variability in disease severity, inflammation sites, and medical therapeutics, may have influenced metabolite detection, as many studies did not stratify subjects based on disease severity. Future research is required to distinguish cause from effect in CD-associated metabolites and examine how CD progression interacts with host and microbial metabolism. Additionally, the limited publicly available data restricts comprehensive analyses, and more data in standardized repositories would enable better assessment and validation of metabolic signatures in Crohn’s disease. Our systematic review identified a comprehensive profile of metabolites that are important in pathophysiology of Crohn’s disease.

## Data Availability

All data produced in the present study are available upon reasonable request to the authors

## Author Contributions

AD, SK, JJR and ACK independently searched studies with the help of MGVN, who helped as a systematic review librarian in retrieving the studies. AD and SK independently extracted the data from the studies. AD performed data analysis and wrote the manuscript with the help of JJR, ACK and MD. SHA, and APA served as peer reviewers and experts in the field to contribute edits and suggestions throughout the writing process. MD conceptualized and designed the study and provided supervision.

## Acknowledgments

This work was supported by UC Davis School of Medicine TriP program (MD and APA). The funding agencies had no role in the study analysis or writing of the manuscript. Its contents are solely the responsibility of the authors.

## References

1 Neurath MF, Travis SP. Mucosal healing in inflammatory bowel diseases: a systematic review. Gut 2012;61:1619–35.

2 Chang JT. Pathophysiology of Inflammatory Bowel Diseases. N Engl J Med 2020;383:2652–64.

3 Dave M, Papadakis KA, Faubion WA, Jr. Immunology of inflammatory bowel disease and molecular targets for biologics. Gastroenterol Clin North Am 2014;43:405–24.

4 Imhann F, Vich Vila A, Bonder MJ, Fu J, Gevers D, Visschedijk MC, et al. Interplay of host genetics and gut microbiota underlying the onset and clinical presentation of inflammatory bowel disease. Gut 2018;67:108–19.

5 Nicholson JK, Lindon JC. Systems biology: Metabonomics. Nature 2008;455:1054–6.

6 Macfarlane GT, Macfarlane S. Fermentation in the human large intestine: its physiologic consequences and the potential contribution of prebiotics. J Clin Gastroenterol 2011;45 **S**uppl:S120-7.

7 Goncalves P, Martel F. Butyrate and colorectal cancer: the role of butyrate transport. Curr Drug Metab 2013;14:994–1008.

8 Cao Y, Shen J, Ran ZH. Association between Faecalibacterium prausnitzii Reduction and Inflammatory Bowel Disease: A Meta-Analysis and Systematic Review of the Literature. Gastroenterol Res Pract 2014;2014:872725.

9 Siddiqui MT, Cresci GAM. The Immunomodulatory Functions of Butyrate. J Inflamm Res 2021;14:6025–41.

10 Vaughn BP, Vatanen T, Allegretti JR, Bai A, Xavier RJ, Korzenik J, et al. Increased Intestinal Microbial Diversity Following Fecal Microbiota Transplant for Active Crohn’s Disease. Inflamm Bowel Dis 2016;22:2182–90.

11 Qin F, Li J, Mao T, Feng S, Li J, Lai M. 2 Hydroxybutyric Acid-Producing Bacteria in Gut Microbiome and Fusobacterium nucleatum Regulates 2 Hydroxybutyric Acid Level In Vivo. Metabolites 2023;13.

12 Adegbola SO, Sarafian M, Sahnan K, Ding NS, Faiz OD, Warusavitarne J, et al. Differences in amino acid and lipid metabolism distinguish Crohn’s from idiopathic/cryptoglandular perianal fistulas by tissue metabonomic profiling and may offer clues to underlying pathogenesis. Eur J Gastroenterol Hepatol 2021;33:1469–79.

13 Nystrom N, Prast-Nielsen S, Correia M, Globisch D, Engstrand L, Schuppe-Koistinen I, et al. Mucosal and Plasma Metabolomes in New-onset Paediatric Inflammatory Bowel Disease: Correlations with Disease Characteristics and Plasma Inflammation Protein Markers. J Crohns Colitis 2023;17:418–32.

14 Costea I, Mack DR, Israel D, Morgan K, Krupoves A, Seidman E, et al. Genes involved in the metabolism of poly-unsaturated fatty-acids (PUFA) and risk for Crohn’s disease in children & young adults. PLoS One 2010;5:e15672.

15 Costea I, Mack DR, Lemaitre RN, Israel D, Marcil V, Ahmad A, et al. Interactions between the dietary polyunsaturated fatty acid ratio and genetic factors determine susceptibility to pediatric Crohn’s disease. Gastroenterology 2014;146:929–31.

16 Lattka E, Illig T, Heinrich J, Koletzko B. Do FADS genotypes enhance our knowledge about fatty acid related phenotypes? Clin Nutr 2010;29:277–87.

17 Liu R, Qiao S, Shen W, Liu Y, Lu Y, Liangyu H, et al. Disturbance of Fatty Acid Desaturation Mediated by FADS2 in Mesenteric Adipocytes Contributes to Chronic Inflammation of Crohn’s Disease. J Crohns Colitis 2020;14:1581–99.

18 Chan SS, Luben R, Olsen A, Tjonneland A, Kaaks R, Lindgren S, et al. Association between high dietary intake of the n-3 polyunsaturated fatty acid docosahexaenoic acid and reduced risk of Crohn’s disease. Aliment Pharmacol Ther 2014;39:834–42.

19 Shoda R, Matsueda K, Yamato S, Umeda N. Epidemiologic analysis of Crohn disease in Japan: increased dietary intake of n-6 polyunsaturated fatty acids and animal protein relates to the increased incidence of Crohn disease in Japan. Am J Clin Nutr 1996;63:741–5.

20 Peters V, Bolte L, Schuttert EM, Andreu-Sanchez S, Dijkstra G, Weersma RK, et al. Western and Carnivorous Dietary Patterns are Associated with Greater Likelihood of IBD Development in a Large Prospective Population-based Cohort. J Crohns Colitis 2022;16:931–9.

21 Li T, Qiu Y, Yang HS, Li MY, Zhuang XJ, Zhang SH, et al. Systematic review and meta-analysis: Association of a pre-illness Western dietary pattern with the risk of developing inflammatory bowel disease. J Dig Dis 2020;21:362–71.

22 Lands B, Bibus D, Stark KD. Dynamic interactions of n-3 and n-6 fatty acid nutrients. Prostaglandins Leukot Essent Fatty Acids 2018;136:15–21.

23 Maceyka M, Spiegel S. Sphingolipid metabolites in inflammatory disease. Nature 2014;510:58–67.

24 Hannun YA, Obeid LM. Principles of bioactive lipid signalling: lessons from sphingolipids. Nat Rev Mol Cell Biol 2008;9:139–50.

25 Spate U, Schulze PC. Proinflammatory cytokines and skeletal muscle. Curr Opin Clin Nutr Metab Care 2004;7:265–9.

26 Yang L, Chu Z, Liu M, Zou Q, Li J, Liu Q, et al. Amino acid metabolism in immune cells: essential regulators of the effector functions, and promising opportunities to enhance cancer immunotherapy. J Hematol Oncol 2023;16:59.

27 Kim MH, Kim H. The Roles of Glutamine in the Intestine and Its Implication in Intestinal Diseases. Int J Mol Sci 2017;18.

28 Felig P, Pozefsky T, Marliss E, Cahill GF, Jr. Alanine: key role in gluconeogenesis. Science 1970;167:1003–4.

29 Vester-Andersen MK, Mirsepasi-Lauridsen HC, Prosberg MV, Mortensen CO, Trager C, Skovsen K, et al. Increased abundance of proteobacteria in aggressive Crohn’s disease seven years after diagnosis. Sci Rep 2019;9:13473.

30 Hey P, Gow P, Testro AG, Apostolov R, Chapman B, Sinclair M. Nutraceuticals for the treatment of sarcopenia in chronic liver disease. Clin Nutr ESPEN 2021;41:13–22.

31 Wang S, van Schooten FJ, Jin H, Jonkers D, Godschalk R. The Involvement of Intestinal Tryptophan Metabolism in Inflammatory Bowel Disease Identified by a Meta-Analysis of the Transcriptome and a Systematic Review of the Metabolome. Nutrients 2023;15.

32 Fernstrom JD, Fernstrom MH. Tyrosine, phenylalanine, and catecholamine synthesis and function in the brain. J Nutr 2007;137:1539S–47S; discussion 48S.

33 Flydal MI, Martinez A. Phenylalanine hydroxylase: function, structure, and regulation. IUBMB Life 2013;65:341–9.

34 Fleishman JS, Kumar S. Bile acid metabolism and signaling in health and disease: molecular mechanisms and therapeutic targets. Signal Transduct Target Ther 2024;9:97.

35 Di Ciaula A, Garruti G, Lunardi Baccetto R, Molina-Molina E, Bonfrate L, Wang DQ, et al. Bile Acid Physiology. Ann Hepatol 2017;16:s4–s14.

36 Ovadia C, Perdones-Montero A, Spagou K, Smith A, Sarafian MH, Gomez-Romero M, et al. Enhanced Microbial Bile Acid Deconjugation and Impaired Ileal Uptake in Pregnancy Repress Intestinal Regulation of Bile Acid Synthesis. Hepatology 2019;70:276–93.

37 Becker HEF, Jamin C, Bervoets L, Boleij A, Xu P, Pierik MJ, et al. Higher Prevalence of Bacteroides fragilis in Crohn’s Disease Exacerbations and Strain-Dependent Increase of Epithelial Resistance. Front Microbiol 2021;12:598232.

38 Kowalska-Duplaga K, Gosiewski T, Kapusta P, Sroka-Oleksiak A, Wedrychowicz A, Pieczarkowski S, et al. Differences in the intestinal microbiome of healthy children and patients with newly diagnosed Crohn’s disease. Sci Rep 2019;9:18880.

39 Hojo A, Kobayashi T, Matsubayashi M, Morikubo H, Miyatani Y, Fukuda T, et al. Usefulness of colestimide for diarrhea in postoperative Crohn’s disease. JGH Open 2022;6:547–53.

40 Choudhuri S, Klaassen CD. Molecular Regulation of Bile Acid Homeostasis. Drug Metab Dispos 2022;50:425–55.

41 Chiang JY. Bile acid metabolism and signaling. Compr Physiol 2013;3:1191–212.

42 Wang YD, Chen WD, Moore DD, Huang W. FXR: a metabolic regulator and cell protector. Cell Res 2008;18:1087–95.

43 Anderson KM, Gayer CP. The Pathophysiology of Farnesoid X Receptor (FXR) in the GI Tract: Inflammation, Barrier Function and Innate Immunity. Cells 2021;10.

44 Chiang JY, Pathak P, Liu H, Donepudi A, Ferrell J, Boehme S. Intestinal Farnesoid X Receptor and Takeda G Protein Couple Receptor 5 Signaling in Metabolic Regulation. Dig Dis 2017;35:241–5.

45 Yoneno K, Hisamatsu T, Shimamura K, Kamada N, Ichikawa R, Kitazume MT, et al. TGR5 signalling inhibits the production of pro-inflammatory cytokines by in vitro differentiated inflammatory and intestinal macrophages in Crohn’s disease. Immunology 2013;139:19–29.

46 Zhou C, Wang Y, Li C, Xie Z, Dai L. Amelioration of Colitis by a Gut Bacterial Consortium Producing Anti-Inflammatory Secondary Bile Acids. Microbiol Spectr 2023;11:e0333022.

47 Jha AK, Huang SC, Sergushichev A, Lampropoulou V, Ivanova Y, Loginicheva E, et al. Network integration of parallel metabolic and transcriptional data reveals metabolic modules that regulate macrophage polarization. Immunity 2015;42:419–30.

48 Schicho R, Shaykhutdinov R, Ngo J, Nazyrova A, Schneider C, Panaccione R, et al. Quantitative metabolomic profiling of serum, plasma, and urine by (1)H NMR spectroscopy discriminates between patients with inflammatory bowel disease and healthy individuals. J Proteome Res 2012;11:3344–57.

49 Yoshida Y, Umeno A, Shichiri M. Lipid peroxidation biomarkers for evaluating oxidative stress and assessing antioxidant capacity in vivo. J Clin Biochem Nutr 2013;52:9–16.

50 Vancamelbeke M, Sabino, J., Deroover, L., Vandermeulen, G., Luypaerts, A., Ferrante, M., Falony, G., Vieira-Silva, S., Verbeke, K., Raes, J., Cleynen, I., & Vermeire, S. Tu1862-Profiling of the fecal microbiota and metabolome in patients with inflammatory bowel disease and their unaffected relatives. Gastroenterology 2017:S991.

51 Dryahina K, Smith D, Bortlik M, Machkova N, Lukas M, Spanel P. Pentane and other volatile organic compounds, including carboxylic acids, in the exhaled breath of patients with Crohn’s disease and ulcerative colitis. J Breath Res 2017;12:016002.

52 Zhgun ES, Kislun YV, Kalachniuk TN, Veselovsky VA, Urban AS, Tikhonova PO, et al. [Evaluation of metabolites levels in feces of patients with inflammatory bowel diseases]. Biomed Khim 2020;66:233–40.

53 Marchesi JR, Holmes E, Khan F, Kochhar S, Scanlan P, Shanahan F, et al. Rapid and noninvasive metabonomic characterization of inflammatory bowel disease. J Proteome Res 2007;6:546–51.

54 Alonso A, Julia A, Vinaixa M, Domenech E, Fernandez-Nebro A, Canete JD, et al. Urine metabolome profiling of immune-mediated inflammatory diseases. BMC Med 2016;14:133.

55 Bjerrum JT, Wang Y, Hao F, Coskun M, Ludwig C, Gunther U, et al. Metabonomics of human fecal extracts characterize ulcerative colitis, Crohn’s disease and healthy individuals. Metabolomics 2015;11:122–33.

56 Hu J, Cheng S, Yao J, Lin X, Li Y, Wang W, et al. Correlation between altered gut microbiota and elevated inflammation markers in patients with Crohn’s disease. Front Immunol 2022;13:947313.

57 Lloyd-Price J, Arze C, Ananthakrishnan AN, Schirmer M, Avila-Pacheco J, Poon TW, et al. Multi-omics of the gut microbial ecosystem in inflammatory bowel diseases. Nature 2019;569:655–62.

58 Ambrozkiewicz F, Karczmarski J, Kulecka M, Paziewska A, Niemira M, Zeber-Lubecka N, et al. In search for interplay between stool microRNAs, microbiota and short chain fatty acids in Crohn’s disease - a preliminary study. BMC Gastroenterol 2020;20:307.

59 Walton C, Fowler DP, Turner C, Jia W, Whitehead RN, Griffiths L, et al. Analysis of volatile organic compounds of bacterial origin in chronic gastrointestinal diseases. Inflamm Bowel Dis 2013;19:2069–78.

60 De Preter V, Machiels K, Joossens M, Arijs I, Matthys C, Vermeire S, et al. Faecal metabolite profiling identifies medium-chain fatty acids as discriminating compounds in IBD. Gut 2015;64:447–58.

61 Aldars-Garcia L, Gil-Redondo R, Embade N, Riestra S, Rivero M, Gutierrez A, et al. Serum and Urine Metabolomic Profiling of Newly Diagnosed Treatment-Naive Inflammatory Bowel Disease Patients. Inflamm Bowel Dis 2024;30:167–82.

62 Murgia A, Hinz C, Liggi S, Denes J, Hall Z, West J, et al. Italian cohort of patients affected by inflammatory bowel disease is characterised by variation in glycerophospholipid, free fatty acids and amino acid levels. Metabolomics 2018;14:140.

63 Di Giovanni N, Meuwis MA, Louis E, Focant JF. Untargeted Serum Metabolic Profiling by Comprehensive Two-Dimensional Gas Chromatography-High-Resolution Time-of-Flight Mass Spectrometry. J Proteome Res 2020;19:1013–28.

64 Esteve-Comas M, Ramirez M, Fernandez-Banares F, Abad-Lacruz A, Gil A, Cabre E, et al. Plasma polyunsaturated fatty acid pattern in active inflammatory bowel disease. Gut 1992;33:1365–9.

65 Figler M, Gasztonyi B, Cseh J, Horvath G, Kisbenedek AG, Bokor S, et al. Association of n-3 and n-6 long-chain polyunsaturated fatty acids in plasma lipid classes with inflammatory bowel diseases. Br J Nutr 2007;97:1154–61.

66 Jansson J, Willing B, Lucio M, Fekete A, Dicksved J, Halfvarson J, et al. Metabolomics reveals metabolic biomarkers of Crohn’s disease. PLoS One 2009;4:e6386.

67 Kuroki F, Iida M, Matsumoto T, Aoyagi K, Kanamoto K, Fujishima M. Serum n3 polyunsaturated fatty acids are depleted in Crohn’s disease. Dig Dis Sci 1997;42:1137–41.

68 Geerling BJ, v Houwelingen AC, Badart-Smook A, Stockbrugger RW, Brummer RJ. Fat intake and fatty acid profile in plasma phospholipids and adipose tissue in patients with Crohn’s disease, compared with controls. Am J Gastroenterol 1999;94:410–7.

69 Jiang J, Chen L, Sun R, Yu T, Jiang S, Chen H. Characterization of serum polyunsaturated fatty acid profile in patients with inflammatory bowel disease. Ther Adv Chronic Dis 2023;14:20406223231156826.

70 Scoville EA, Allaman MM, Adams DW, Motley AK, Peyton SC, Ferguson SL, et al. Serum Polyunsaturated Fatty Acids Correlate with Serum Cytokines and Clinical Disease Activity in Crohn’s Disease. Sci Rep 2019;9:2882.

71 Akazawa Y, Morisaki T, Fukuda H, Norimatsu K, Shiota J, Hashiguchi K, et al. Significance of serum palmitoleic acid levels in inflammatory bowel disease. Sci Rep 2021;11:16260.

72 Kruchinina MV, Gromov, A. A., Svetlova, I. O., Azgaldyan, A. V., Kruchinin, V. N., Shashkov, M. V., & Sokolova, A. S. P0388-New diagnostic capabilities for crohn’s disease: Use of fatty acid profiles of erythrocyte membranes and blood serum. United European Gastroenterology Journal 2020 334–5.

73 Ben-Mustapha Y, Ben-Fradj MK, Hadj-Taieb S, Serghini M, Ben Ahmed M, Boubaker J, et al. Altered mucosal and plasma polyunsaturated fatty acids, oxylipins, and endocannabinoids profiles in Crohn’s disease. Prostaglandins Other Lipid Mediat 2023;168:106741.

74 Lai Y, Xue J, Liu CW, Gao B, Chi L, Tu P, et al. Serum Metabolomics Identifies Altered Bioenergetics, Signaling Cascades in Parallel with Exposome Markers in Crohn’s Disease. Molecules 2019;24.

75 Levy E, Rizwan Y, Thibault L, Lepage G, Brunet S, Bouthillier L, et al. Altered lipid profile, lipoprotein composition, and oxidant and antioxidant status in pediatric Crohn disease. Am J Clin Nutr 2000;71:807–15.

76 Alghamdi A, Gerasimidis K, Blackburn G, Akinci D, Edwards C, Russell RK, et al. Untargeted Metabolomics of Extracts from Faecal Samples Demonstrates Distinct Differences between Paediatric Crohn’s Disease Patients and Healthy Controls but No Significant Changes Resulting from Exclusive Enteral Nutrition Treatment. Metabolites 2018;8.

77 Fathi F, Majari-Kasmaee L, Mani-Varnosfaderani A, Kyani A, Rostami-Nejad M, Sohrabzadeh K, et al. 1H NMR based metabolic profiling in Crohn’s disease by random forest methodology. Magn Reson Chem 2014;52:370–6.

78 Kruchinina MV, Gromov, A. A., Svetlova, I. O., Azgaldyan, A. V., Kruchinin, V. N., Shashkov, M. V., Sokolova, A. S., Yakovina, I. N., & Shestov, A. A. . P0305-Peculiarities of fatty acid profiles of erythrocyte membranes and blood serum depending on activity rate of Crohn’s disease. United European Gastroenterology Journal 2021:436.

79 Franzosa EA, Sirota-Madi A, Avila-Pacheco J, Fornelos N, Haiser HJ, Reinker S, et al. Gut microbiome structure and metabolic activity in inflammatory bowel disease. Nat Microbiol 2019;4:293–305.

80 Chen B, Wang Y, Wang Q, Li D, Huang X, Kuang X, et al. Untargeted metabolomics identifies potential serum biomarkers associated with Crohn’s disease. Clin Exp Med 2023;23:1751–61.

81 Filimoniuk A, Blachnio-Zabielska A, Imierska M, Lebensztejn DM, Daniluk U. Sphingolipid Analysis Indicate Lactosylceramide as a Potential Biomarker of Inflammatory Bowel Disease in Children. Biomolecules 2020;10.

82 Braun A, Treede I, Gotthardt D, Tietje A, Zahn A, Ruhwald R, et al. Alterations of phospholipid concentration and species composition of the intestinal mucus barrier in ulcerative colitis: a clue to pathogenesis. Inflamm Bowel Dis 2009;15:1705–20.

83 Ferru-Clement R, Boucher G, Forest A, Bouchard B, Bitton A, Lesage S, et al. Serum Lipidomic Screen Identifies Key Metabolites, Pathways, and Disease Classifiers in Crohn’s Disease. Inflamm Bowel Dis 2023;29:1024–37.

84 Fan F, Mundra PA, Fang L, Galvin A, Moore XL, Weir JM, et al. Lipidomic Profiling in Inflammatory Bowel Disease: Comparison Between Ulcerative Colitis and Crohn’s Disease. Inflamm Bowel Dis 2015;21:1511–8.

85 Iwatani S, Iijima H, Otake Y, Amano T, Tani M, Yoshihara T, et al. Novel mass spectrometry-based comprehensive lipidomic analysis of plasma from patients with inflammatory bowel disease. J Gastroenterol Hepatol 2020;35:1355–64.

86 Kurada S, Grove, D., Mao, R., Singh, A., Sarvepalli, S., Lopez, R., Fiocchi, C., Rogler, G., Kunst, C., Dweik, R., & Rieder, F. . Sa1789-Serum headspace metabolome analysis discriminates crohn’s disease from controls with high accuracy and is linked to disease phenotypes. Gastroenterology 2018 S-395.

87 Williams HR, Willsmore JD, Cox IJ, Walker DG, Cobbold JF, Taylor-Robinson SD, et al. Serum metabolic profiling in inflammatory bowel disease. Dig Dis Sci 2012;57:2157–65.

88 Jagt JZ, Struys EA, Ayada I, Bakkali A, Jansen EEW, Claesen J, et al. Fecal Amino Acid Analysis in Newly Diagnosed Pediatric Inflammatory Bowel Disease: A Multicenter Case-Control Study. Inflamm Bowel Dis 2022;28:755–63.

89 Powles STR, Gallagher, K., Hicks, L. C., Chong, L. W., Swann, J. R., Holmes, E., Williams, H. R. T., & Orchard, T. R. PTH-112 Effect of co-morbidities in crohn’s disease associated urinary metabolic profiles. Gut 2019:A89–A90.

90 Lins Neto MAF, Verdi GMX, Veras AO, Veras MO, Caetano LC, Ursulino JS. Use of Metabolomics to the Diagnosis of Inflammatory Bowel Disease. Arq Gastroenterol 2020;57:311–5.

91 Ooi M, Nishiumi S, Yoshie T, Shiomi Y, Kohashi M, Fukunaga K, et al. GC/MS-based profiling of amino acids and TCA cycle-related molecules in ulcerative colitis. Inflamm Res 2011;60:831–40.

92 Scoville EA, Allaman MM, Brown CT, Motley AK, Horst SN, Williams CS, et al. Alterations in Lipid, Amino Acid, and Energy Metabolism Distinguish Crohn’s Disease from Ulcerative Colitis and Control Subjects by Serum Metabolomic Profiling. Metabolomics 2018;14:17.

93 Kolho KL, Pessia A, Jaakkola T, de Vos WM, Velagapudi V. Faecal and Serum Metabolomics in Paediatric Inflammatory Bowel Disease. J Crohns Colitis 2017;11:321–34.

94 Krzystek-Korpacka M, Fleszar MG, Bednarz-Misa I, Lewandowski L, Szczuka I, Kempinski R, et al. Transcriptional and Metabolomic Analysis of L-Arginine/Nitric Oxide Pathway in Inflammatory Bowel Disease and Its Association with Local Inflammatory and Angiogenic Response: Preliminary Findings. Int J Mol Sci 2020;21.

95 Owczarek D, Cibor D, Mach T. Asymmetric dimethylarginine (ADMA), symmetric dimethylarginine (SDMA), arginine, and 8-iso-prostaglandin F2alpha (8-iso-PGF2alpha) level in patients with inflammatory bowel diseases. Inflamm Bowel Dis 2010;16:52–7.

96 Herlekar D, Martin, F. P., Zhang, S., Bosworth, B., Montoliu, I., Collino, S., Yantiss, R., Kochnar, S., Scherl, E., Dogan, B., & Simpson, K. Chemical perturbations in the inflamed ileum promote the growth of crohn’s associated adherent and invasive E. coli. Inflammatory Bowel Diseases 2013:S121.

97 Tews HC, Schmelter F, Kandulski A, Buchler C, Schmid S, Schlosser S, et al. Unique Metabolomic and Lipidomic Profile in Serum From Patients With Crohn’s Disease and Ulcerative Colitis Compared With Healthy Control Individuals. Inflamm Bowel Dis 2023.

98 Williams HR, Cox IJ, Walker DG, North BV, Patel VM, Marshall SE, et al. Characterization of inflammatory bowel disease with urinary metabolic profiling. Am J Gastroenterol 2009;104:1435–44.

99 Bosch S, Struys EA, van Gaal N, Bakkali A, Jansen EW, Diederen K, et al. Fecal Amino Acid Analysis Can Discriminate De Novo Treatment-Naive Pediatric Inflammatory Bowel Disease From Controls. J Pediatr Gastroenterol Nutr 2018;66:773–8.

100 Levhar N, Hadar, R., Braun, T., Efroni, G., Abramovich, I., Gottlieb, E., Sosnovski, K., Abass, H., Berger, T., Granot, M., Neuman, S., Selinger Feigin, L., Picard, O., Yavzuri, M., Lahat, A., Eliakim, A., Weiss, B., Kopylov, U., Ben-Horin, S., Amir, A., Haberman, Y. . P0239-Untargeted serum metabolome in longitudinal Crohn’s disease (CD) cohort enrolled during remission shows strong individualized signature and CD-associated signals that are maintained also in patients who normalized their fecal calprotectin. United European Gastroenterology Journal 2021 396.

101 Morasso C, Truffi M, Vanna R, Albasini S, Mazzucchelli S, Colombo F, et al. Raman Analysis Reveals Biochemical Differences in Plasma of Crohn’s Disease Patients. J Crohns Colitis 2020;14:1572–80.

102 Gupta NK, Thaker AI, Kanuri N, Riehl TE, Rowley CW, Stenson WF, et al. Serum analysis of tryptophan catabolism pathway: correlation with Crohn’s disease activity. Inflamm Bowel Dis 2012;18:1214–20.

103 Nikolaus S, Schulte B, Al-Massad N, Thieme F, Schulte DM, Bethge J, et al. Increased Tryptophan Metabolism Is Associated With Activity of Inflammatory Bowel Diseases. Gastroenterology 2017;153:1504–16 e2.

104 Conrad M, Bittinger, K., Ren, Y., Dawany, N., Baldassano, R., Bushman, F., & Kelsen, J. Taxonomic and metabolic differences among very early onset inflammatory bowel disease, older onset pediatric inflammatory bowel disease. Journal of Pediatric Gastroenterology and Nutrition 2018:S197–S8.

105 Gnewuch C, Liebisch G, Langmann T, Dieplinger B, Mueller T, Haltmayer M, et al. Serum bile acid profiling reflects enterohepatic detoxification state and intestinal barrier function in inflammatory bowel disease. World J Gastroenterol 2009;15:3134–41.

106 Jacobs JP, Goudarzi M, Singh N, Tong M, McHardy IH, Ruegger P, et al. A Disease-Associated Microbial and Metabolomics State in Relatives of Pediatric Inflammatory Bowel Disease Patients. Cell Mol Gastroenterol Hepatol 2016;2:750–66.

107 Weng YJ, Gan HY, Li X, Huang Y, Li ZC, Deng HM, et al. Correlation of diet, microbiota and metabolite networks in inflammatory bowel disease. J Dig Dis 2019;20:447–59.

108 Sun R, Jiang J, Yang L, Chen L, Chen H. Alterations of Serum Bile Acid Profile in Patients with Crohn’s Disease. Gastroenterol Res Pract 2022;2022:1680008.

109 Feng L, Zhou N, Li Z, Fu D, Guo Y, Gao X, et al. Co-occurrence of gut microbiota dysbiosis and bile acid metabolism alteration is associated with psychological disorders in Crohn’s disease. FASEB J 2022;36:e22100.

110 Ju J, Zhang C, Yang J, Yang Q, Yin P, Sun X. Deoxycholic acid exacerbates intestinal inflammation by modulating interleukin-1beta expression and tuft cell proportion in dextran sulfate sodium-induced murine colitis. PeerJ 2023;11:e14842.

111 Kiasat A, Prast-Nielsen S, Rautiainen S, Engstrand L, Andersson F, Lindberg J, et al. Plasma bile acids in association with Crohn’s disease. Scand J Gastroenterol 2024;59:674–82.

112 Lu X, Zhou, M., Qu, C., Ge, W., Chen, Y. Serum Bile Acid Profile in Patients with Inflammatory Bowel Disease. Chinese Journal of Gastroenterology 2017;22.

113 Ma R, Zhu Y, Li X, Hu S, Zheng D, Xiong S, et al. A Novel Serum Metabolomic Panel for the Diagnosis of Crohn’s Disease. Inflamm Bowel Dis 2023;29:1524–35.

114 Murakami M, Iwamoto J, Honda A, Tsuji T, Tamamushi M, Ueda H, et al. Detection of Gut Dysbiosis due to Reduced Clostridium Subcluster XIVa Using the Fecal or Serum Bile Acid Profile. Inflamm Bowel Dis 2018;24:1035–44.

115 Liu H, Xu M, He Q, Wei P, Ke M, Liu S. Untargeted serum metabolomics reveals specific metabolite abnormalities in patients with Crohn’s disease. Front Med (Lausanne) 2022;9:814839.

116 Rieder F, Kurada S, Grove D, Cikach F, Lopez R, Patel N, et al. A Distinct Colon-Derived Breath Metabolome is Associated with Inflammatory Bowel Disease, but not its Complications. Clin Transl Gastroenterol 2016;7:e201.

117 Kurada S, Grove, D., Alkhouri, N., Lopez, R., Brzezinski, A., Baker, M., Fiocchi, C., Dweik, R., & Rieder, F. A specific breath metabolome signature identifies patients with inflammatory bowel diseases. American Journal of Gastroenterology 2015 S783.

118 Hicks L, Walker, D., Eng, D., Jiminez, B., Kinross, J., Holmes, E., Williams, H., & Orchard, T. Urinary metabolic profiling of inflammatory bowel disease in a South Asian cohort Journal of Crohn’s and Colitis 2013:S258–S9.

119 Williams HR, Cox IJ, Walker DG, Cobbold JF, Taylor-Robinson SD, Marshall SE, et al. Differences in gut microbial metabolism are responsible for reduced hippurate synthesis in Crohn’s disease. BMC Gastroenterol 2010;10:108.

120 Bodelier AG, Smolinska A, Baranska A, Dallinga JW, Mujagic Z, Vanhees K, et al. Volatile Organic Compounds in Exhaled Air as Novel Marker for Disease Activity in Crohn’s Disease: A Metabolomic Approach. Inflamm Bowel Dis 2015;21:1776–85.

121 Hrdlicka L, Dryahina, K., Spanel, P., Bortlik, M., Duricova, D., Machkova, N., & Lukas, M. . Noninvasive quantification of volatile metabolites in breath: A potential indicator of inflammatory bowel diseases activity. . Gastroenterology 2012:S784.

122 Dryahina K, Spanel P, Pospisilova V, Sovova K, Hrdlicka L, Machkova N, et al. Quantification of pentane in exhaled breath, a potential biomarker of bowel disease, using selected ion flow tube mass spectrometry. Rapid Commun Mass Spectrom 2013;27:1983–92.

123 Wendland BE, Aghdassi E, Tam C, Carrrier J, Steinhart AH, Wolman SL, et al. Lipid peroxidation and plasma antioxidant micronutrients in Crohn disease. Am J Clin Nutr 2001;74:259–64.

124 Ludek H. Pentane and carbon disulphide breath concentrations: A potential indicator of inflammatory bowel diseases activity. Journal of Gastrointestinal and Liver Diseases 2012: 21.

