## supplementary information for "Metabolic alterations in Crohn’s disease: A Systematic Review"

**Conflict of Interest Disclosures:** SHA is founder and principal of XenoMed LLC, which has interest in microbial metabolism; however, XenoMed had no role in the analyses or writing of the manuscript.

**Keywords:** Crohn's disease; metabolomics, metabolites, bile acids, lipids and fatty acids, biogenic amines, gut dysbiosis.

### Methods

**Study selection:** Study selection was made on the following inclusion criteria: (1) Human studies, (2) Study included minimum 10 CD patients and non-IBD controls, (3) Involved metabolites detection in any biological sample including serum, plasma, stool, intestinal tissues or breath, (4) Involved chemistry analytical techniques like chromatography, spectroscopy, mass-spectrometry to quantify the metabolites, (5) published as a peer-review article, letter, or abstract. Exclusion criteria include: (1) non-Human studies, (2) studies not included use of any chemistry analytical techniques, (3) Included <10 CD patients.

**Data Extraction:** Two authors (AD and SK) independently performed data extraction from the selected studies using customized data extraction templates. The template includes: (1) First author of the study and year of publication, (2) Title of the study, (3) Number of non-IBD controls included in the study, (4) Number of CD patients included in the study, (5) Type of biological samples used in the study, (6) Chemistry analytical technique used in the study, (7) Metabolites decreased in CD patients compared to non-CD controls, (8) Metabolites elevated in CD patients compared to non-CD controls.

**Data analysis**: Only annotated (named) metabolites were compared while analyzing the dysregulation pattern. Thes metabolites were categorized into lipids and fatty acids, biogenic amines and peptides, bile acids and bile salts, and other primary metabolites. Relative levels of metabolites are only reported in the results section if the change were statistically significant or identified as a discriminating variable when comparing CD patients and non-IBD controls in two or more studies.

**Figure S1**. Metabolomics identifies metabolites that are varied in chemical diversity and implicated in biological phenotypes and disease states.

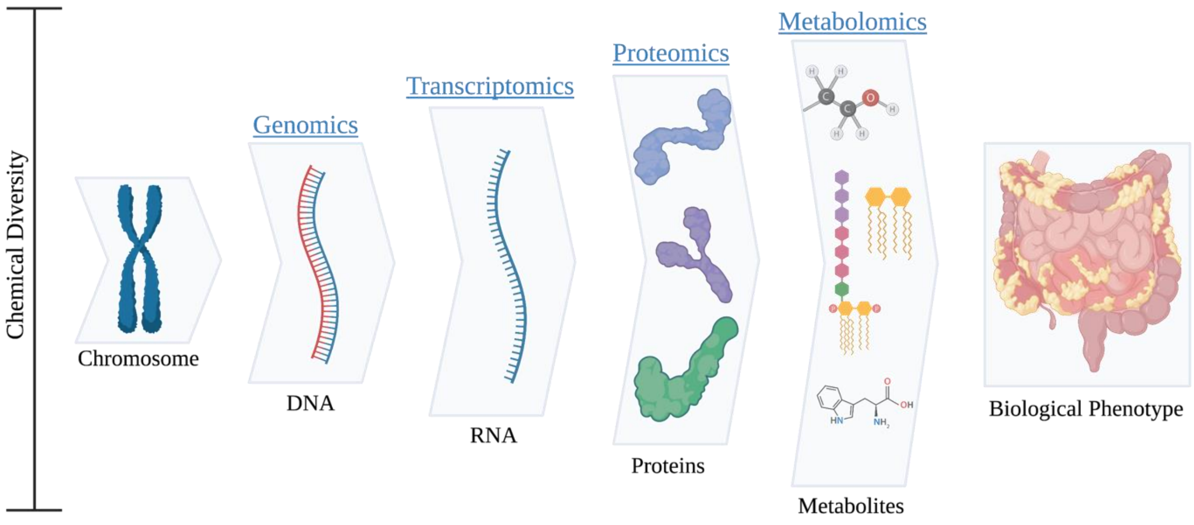

**Figure S2.** Reductions in short chain fatty acids such as butyric acid affect multiple immune pathways implicated in bacterial overgrowth. In terms of dysbiosis, reductions in SCFAs lead to reduced active oxygen consumption, increasing oxygen concentration in the intestine lumen, and promoting aerobic bacteria overgrowth. Reduced SCFAs have also been shown to increase cytokine production, TNFα signaling, and NF-κB-induced gene expression which play important roles in chronic inflammation.

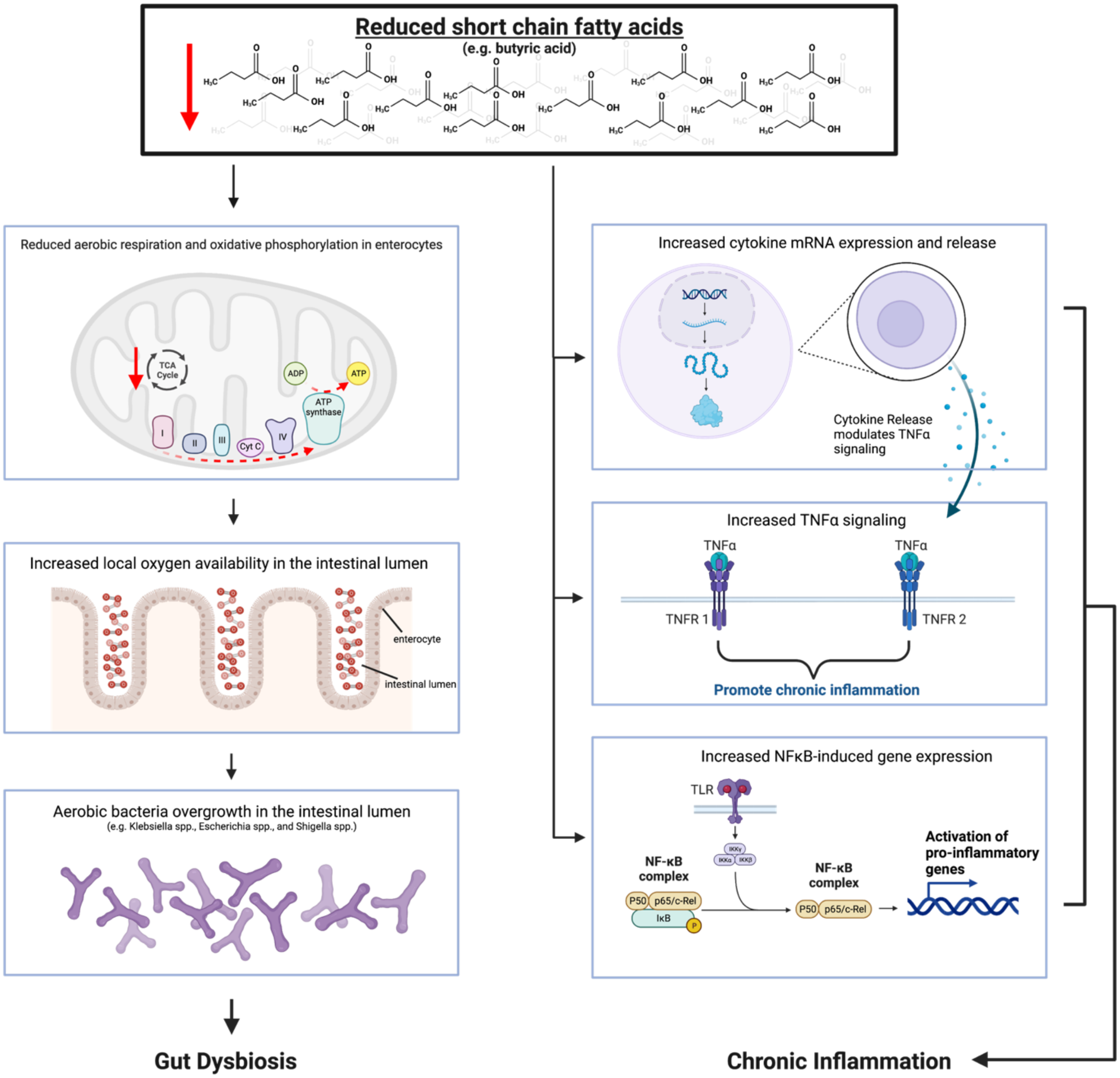

**Figure S3.** Model of dysregulation of ceramides and S1P in CD, leading to increased activity of proinflammatory mediators. Inflammation of colonic enterocytes and dysbiosis with overgrowth of commensals (such as *B. fragilis)* lead to increased number of atrophic intestinal epithelial cell membranes that are shed into the lumen and increase the amount of excreted lipids. This loss of lipids leads to activation of enzymes facilitating ceramide production. Additionally, ceramide derived S1P levels are also increased in this pro-inflammatory state. Ceramides and S1P lead to the direct activation of the associated proinflammatory mediators Il-6 and STAT3 as well as NK cell activation.

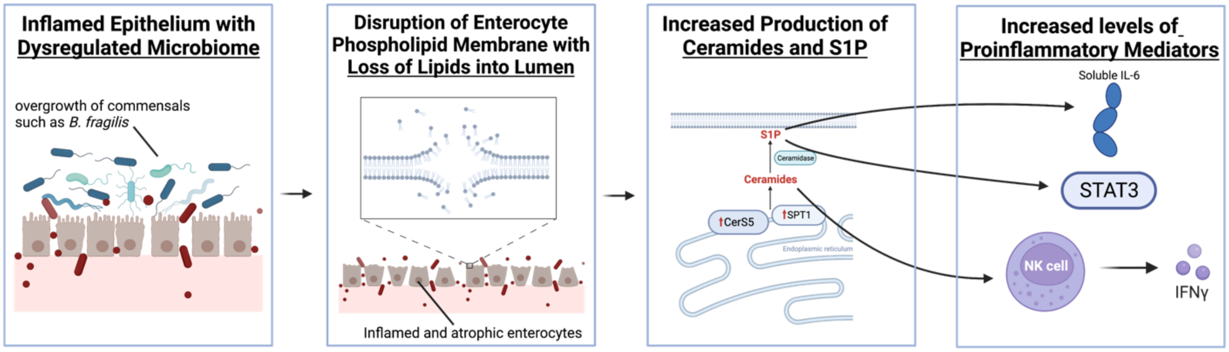

**Figure S4.** Immunomodulatory effects of secondary bile acids. Secondary bile acids activate nuclear farnesoid X receptor (FXR) in enterocytes leading to inhibition of interleukin-1-β and interleukin 8 release. FXR is also expressed in dendritic cells that affect T regulatory cells to diminish their immune-stimulatory properties. Secondary bile acids also activate membrane Takeda G protein-coupled receptor 5 (TGR5) on macrophages to express their anti-inflammatory phenotype as well as on intestinal stem cells to promote growth and differentiation.

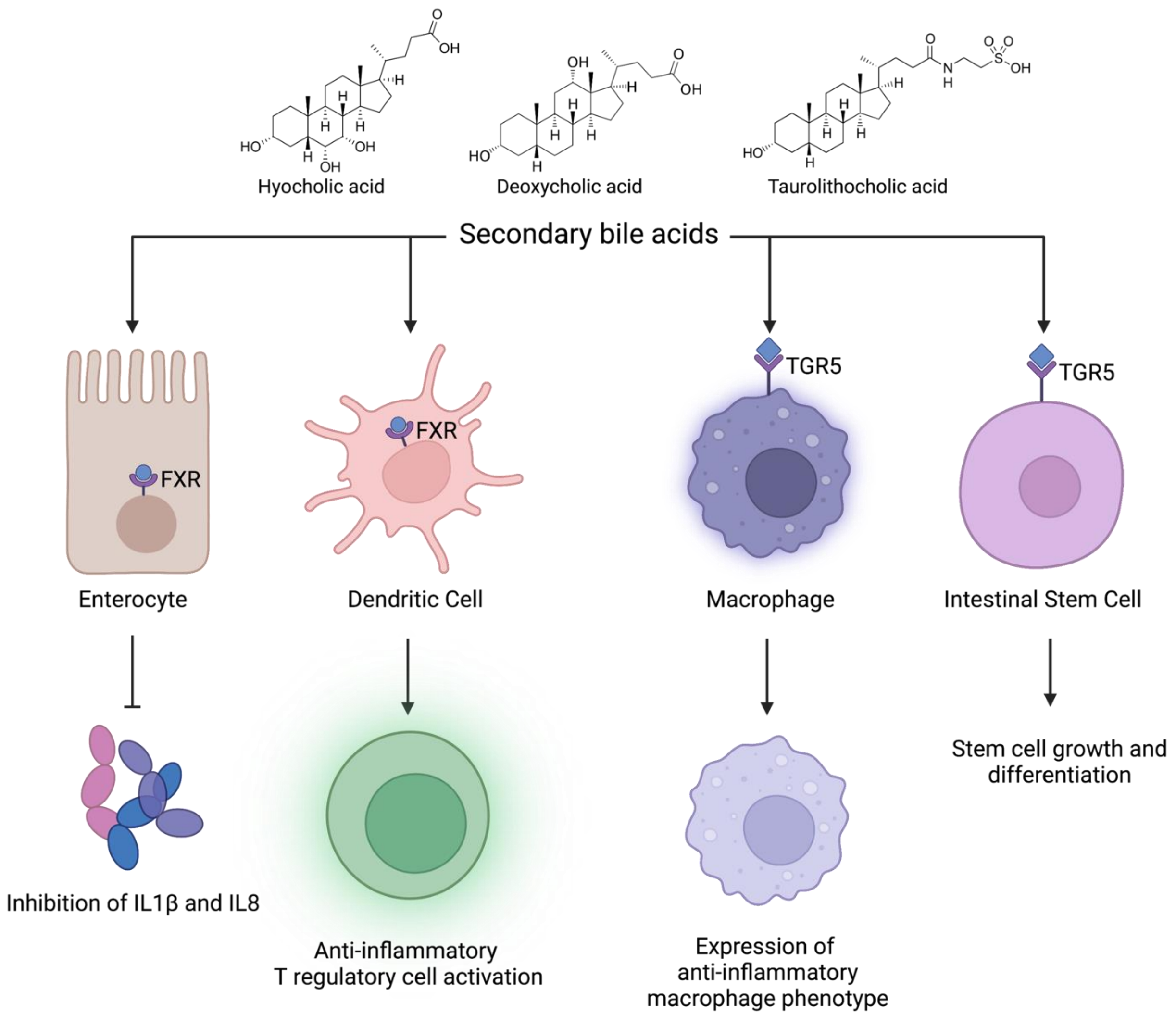

**Supplementary Table**

**Table S1**. MeSH (Medical Subject Headings) terms used for systematic search.

| Category 1: Disease | Category 2: Metabolomic Data | Category 3: High-Throughput Method |
|---|---|---|
| Crohn's Disease | Metabolomics | Chemistry Analytical Techniques |
| Inflammatory Bowel Disease | Metabolome | |
| | Biomarkers | |
| | Lipids | |
| | Fatty acid | |
| | Membrane lipids | |
| | Amino Acids | |
| | Amines | |
| | Carbohydrate | |
| | Bile acids and salts | |
